# Classifying the severity of diabetic macular oedema from optical coherence tomography scans using deep learning: a feasibility study

**DOI:** 10.1101/2025.02.19.24317749

**Authors:** Cathal Breathnach, Fiona Harney, Deirdre Townley, Rachel Hickey, Andrew Simpkin, Derek O’Keeffe

**Affiliations:** School of Medicine, College of Medicine Nursing and Health Sciences, University of Galway, Galway, Ireland; Department of Ophthalmology, University Hospital Galway, Galway, Ireland; School of Mathematical and Statistical Sciences, University of Galway, Galway, Ireland; Department of Endocrinology, University Hospital Galway, Galway, Ireland

## Abstract

**Background:** Diabetic macular oedema (DME) is a vision-threatening complication of diabetes mellitus. It is reliably detected using optical coherence tomography (OCT). This work evaluates a deep learning system (DLS) for the automated detection and classification of DME severity from OCT images.

**Methods:** Anonymised OCT images were retrospectively obtained from 950 patients at University Hospital Galway, Ireland. Images were graded by a consultant ophthalmologist to classify the level of DME present (normal, non-centre-involving DME, centre-involving DME) excluding other pathologies. A DLS was trained using cross-validation, then evaluated on a test dataset and an external dataset. The test set was graded by a second ophthalmologist for comparison.

**Results:** In detecting the presence of DME, the DLS achieved a mean area under the receiver operating characteristic curve (AUC) of 0.98 on cross-validation. AUCs of 0.94 (95% CI 0.90-0.98) and 0.94 (0.92-0.96) were achieved on evaluation of DME detection for the test dataset when graded by the first and second ophthalmologist respectively. An AUC of 0.94 (0.92-0.96) was achieved on evaluation with the external dataset. When detecting the DME severity, AUCs of 0.98, 0.86 and 0.99 were achieved per class on cross validation. For the test dataset, AUCs of 0.99, 0.89 and 0.98 were achieved when graded by the first ophthalmologist and AUCs of 0.96, 0.89 and 0.95 were achieved when graded by the second ophthalmologist.

**Conclusion:** This study suggests promising results for the use of deep learning in the classification of severity of DME which could be used to automate screening for DME and direct appropriate referrals.

## INTRODUCTION

Diabetic macular oedema (DME) is the accumulation of fluid in the macula that can occur at any stage of diabetic retinopathy (DR).[1] DME is associated with more severe DR, and can result in visual loss.[2] Early detection facilitates treatment, including systemic control of diabetes alongside treatments such as laser photocoagulation and intraocular anti-vascular endothelial growth factor (VEGF) injections.[3] Many DR screening programmes use retinal photographs for DME diagnosis, but this relies on surrogate markers, which is associated with high false positive rates, leading to unnecessary referrals.[3–5] In comparison, optical coherence tomography (OCT) provides a detailed structural view of the retina and is the gold standard for DME evaluation.[4,6] OCT is quick, non-invasive and has been recommended for use in screening programmes to reduce unneeded referrals and cost.[4] Certain screening programmes already routinely use OCT for example in Denmark,[7] and in Ireland, where it is used for higher risk patients and has been predicted to reduce referrals.[8] However, screening processes are often manual, and OCT scans require expert interpretation.[2,9]

Deep learning (DL) is a form of artificial intelligence (AI) that can be used to analyse OCT scans.[9,10] A convolutional neural network (CNN) is a class of DL algorithm that is commonly used for medical image classification,[11] and such algorithms have been shown to reliably detect DME from OCT scans and have the potential to play a role in treatment decisions.[12–14] Several approaches for the implementation of DL algorithms for OCT analysis have previously been tested.[15–27] Some studies have analysed two-dimensional images from OCT images (B-scans) for the detection of DME and other retinal pathologies.[15–17] Other studies have used a transfer learning approach (which uses an existing model trained to recognise general images and then optimises it for a supplied dataset) which can offer an advantage when very large datasets are not available.[11] This approach has been tested for the detection of DME and other retinal pathologies such as choroidal neovascularization and drusen.[11,18,19] Several studies have specifically analysed three-dimensional OCT volumes to detect DME alongside age-related macular degeneration.[20–22] Other studies have taken an approach for full segmentation of the retina to diagnose disease, using an additional segmentation step to allow the detection of various pathologies or to segment retinal fluid.[23–25] A notable study was developed by De Fauw et al., using an initial segmentation approach to reliably predict a referral level.[26] However, while each study reports favourable performance for the detection of DME and other retinal pathologies, there are limitations. For example, the segmentation approach requires labour intensive annotation and approaches such as transfer learning can require considerable input to fine-tune the algorithm.

However, few studies including those outlined thus far consider the severity of the DME present, only detecting whether it is present or absent. Various terms have been used to describe DME, including clinically-significant diabetic macular oedema, which can be determined by the presence of specific characteristics.[28] The central area of the fovea is of most interest, and DME within 500μm on either side of the foveal centre is referred to as centrally-involved DME (CI-DME).[2] The International Council of Ophthalmology (ICO) recommends that the presence of CI-DME should be used in conjunction with visual acuity to guide treatment decisions.[2] A study by Tang et al. considered this difference when testing a DL approach for DME detection and classification.[27] This study achieved favourable performance and was tested using both two-dimensional images and three-dimensional scan data. A further more recent study also utilised three dimensional OCT scan data to classify the DME severity per the same classification system, also showing promising results.[29] Additionally, there are specific biomarkers detectable using OCT scans that can be used to define the DME severity,[30] and it has been suggested in a recent review that such biomarkers could be used to direct treatment decisions.[31] A study by Xu et al. used DL to analyse for the presence of such features,[32] and a more recent study by Mitamura et al. also utilised biomarkers combined with visual acuity to guide treatment.[33] These approaches did however require segment annotation, which as outlined previously is time consuming. Tan et al. did also examine the use of deep learning to detect CI-DME with visual impairment from OCT images combined with fundal photographs but showed no benefit over using OCT alone to detect CI-DME.[34] Another recent study by Mondal et al. has used deep learning to predict patient responses to treatments such as anti-VEGF injections, but this study only included patients who had already been diagnosed with DME.[35]

Thus, limited studies have examined the classification of DME from standard OCT images that could be utilised in a screening setting with the exception of the study completed by Tang et al.,[27] and it is clear this area is under-explored. Our study will implement a simple and lightweight approach to this problem, using a recognised scale for DME severity. Furthermore, we are not aware of any study that has previously tested DL for the detection of diabetic eye disease in an Irish population.

## MATERIALS & METHODS

### Image Collection & Datasets

Patients were randomly selected from all patients with diabetes currently being treated in University Hospital Galway under the Irish national DR screening programme to create the training and validation dataset. Imaging was conducted with a Heidelberg Spectralis OCT machine. A volume scan was captured for each patient, but one single B-scan, from the level of the fovea was used as the input for this algorithm. The algorithm was designed for a square input and thus a square image centred around the fovea was used, being cropped if required. Images were taken from both eyes when available. As a retrospective study, there was no patient follow-up and there was no patient or public involvement in the design or conduct of this study.

A second test dataset was created using scan images from every patient in the diabetic registry who attended for an ophthalmology appointment over a two-week period in July 2022. No patients from the training and validation dataset were included in the test dataset. Finally, an externally graded dataset of 250 images, assessed by multiple graders, was also used to test the algorithm.[36] Only DME detection was tested using this external set as these images were not graded using the scheme used in this study.

### Grading Criteria

This study aimed to distinguish between normal OCT images and those with signs of DME, but it also classified the level of DME present. The scale used is as defined by the ICO guidelines for the treatment of DME, using DME changes within the central area, within 500μm of the foveal centre as the cutoff between categories.[2] A description used is outlined in Table 1 with Figure 1 providing image examples as per this scheme. This central zone is the area of most significance and this may become the thickest part of the retina with DME.[28] This grading scheme has a clinical context in mind, as DME that affects the central area poses an increased threat to a patient’s vision and will be more likely to require and respond to DME treatments such as intra-vitreal anti-VEGF injections.[2] This is clinically relevant for a screening programme, where the aim is to detect those who need the most urgent treatment in over-burdened services. Combining the presence of CI-DME with visual acuity may even offer an additional option for triage as while CI-DME may later develop into more substantial DME, it has been shown that it does not always require urgent treatment, and may be suitable for monitoring if visual acuity is not affected.[31] Thus, by choosing this grading scale, patients could be triaged more effectively, as those with more severe disease could be referred for more immediate treatment, while those with other signs of DME could be suitable for further assessment of visual acuity, followed by treatment as required.

**Table 1:** DME classification requirements. *The central area is defined as 500μm on either side of the foveal centre.

| DME Classification | Requirements |
| --- | --- |
| **NORMAL** | No signs of DME. |
| **NCI-DME** | Signs of DME present only outside of the central area*. |
| **CI-DME** | Any signs of DME present within the central area*. |

DME, diabetic macular oedema; NC, non-centre-involving; CI, centre-involving.

**Figure 1:**
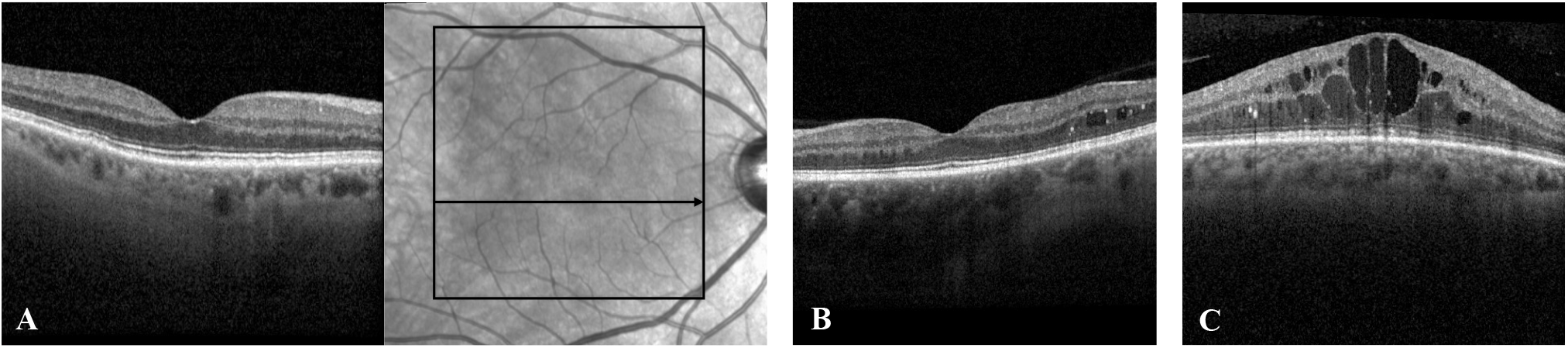
OCT image examples as per the grading scheme used in this study, showing an example of a normal scan with corresponding level on a retinal photograph (A), an example of NCI-DME showing DME with no change to central area (B) and an example of CI-DME with central changes and loss of the normal foveal contour (C).

### Exclusion Criteria

Exclusion criteria were used to exclude non-relevant images for the scope of this study. The study excluded (i) poor quality scans, where the retinal layers at the level of the fovea are not discernible, (ii) images with signs of other retinal pathologies, an example of which is wet age-related macular degeneration, (iii) loss of the foveal contour caused by another retinal pathology, for example due to an epiretinal membrane and (iv) images that did not represent the foveal level.

### Grading Process

The training and validation dataset was graded by an ophthalmology consultant (first ophthalmologist) as per the grading scheme outlined. This ophthalmologist also determined which images met the exclusion criteria for the training and validation data.

The test dataset was graded by the first ophthalmologist and by a second ophthalmology consultant (second ophthalmologist) for comparison. The graders were both aware of the exclusion criteria in this case and there was agreement on which images to exclude. Both graders discussed the grading criteria to reach a consensus but then graded the images independently. Cohen’s k [37] was also calculated as a measure of agreement in grading. The algorithm trained with the training and validation data generated predictions for the test dataset and were compared independently to the grades produced by the two ophthalmologists. Both consultant ophthalmologists involved in this study had more than 10 years of experience in reading OCT scans and treating diabetic eye disease. No clinical details were provided for image grading.

### Algorithm Overview

A CNN was designed and implemented using the Python programming language (version 3.8.13) with the Keras (version 2.9.0) and Tensorflow frameworks (version 2.9.2). The training and validation stage used stratified 5-fold cross validation. The structure of the model is outlined in Supplemental Table 1 and is approximately similar to CNNs used in other DL studies for OCT image analysis.[15] The overall structure and sizes used were determined experimentally to optimise performance within the scope of this study. Two near-identical models were trained in this study with only a differing output layer. The first model for DME detection, used the sigmoid activation for two-class classification with a threshold of 0.5 and the second model used the softmax activation function for three-class classification of the level of DME. Each convolution layer used zero padding with a relu activation function. The model was trained for 35 epochs in each setting with a learning rate of 0.001, using the Adam optimiser.[38] Data augmentation was used as has been implemented in similar studies,[27] using the Keras library to reduce overfitting. This involved random flipping, 10% random rotation and 10% random zoom applied to the data before the rescaling step.

The performance of the algorithm was measured by reporting accuracy, sensitivity, specificity, positive predictive value (PPV), negative predictive value (NPV) and area under the receiver operating characteristic curve (AUC). Confidence intervals for the performance indices were calculated in the test data using the DeLong method for AUC[39] (*pROC* R package, version 1.18.5) and the Wilson method otherwise [40] using the *epiR* R package (version 2.0.77).

## RESULTS

### Population

Within the available timeframe, a total of 1878 images were included from 950 distinct patients. The training and validation dataset was comprised of 1598 images from 807 patients, and after the exclusion criteria were applied, there were 1501 images available from 793 patients. The test dataset was comprised of 280 images from 143 patients, and after the exclusion criteria were applied, there were 239 images included from 132 patients. The class breakdown is similar between the training and test datasets and is presented with other dataset characteristics in Table 2, graded as per the first ophthalmologist. The test dataset was graded by both the first and second ophthalmologist, and Cohen’s κ was calculated as 0.87 for DME detection and 0.85 for DME classification suggesting near-perfect agreement in both situations.

**Table 2:** Characteristics of the training and test data collected for all non-excluded images. Percentage values expressed relative to the training or test datasets. Graded as per the first ophthalmologist.

| Characteristic | Training Dataset |  | Test Dataset |  |
| --- | --- | --- | --- | --- |
|  | n | % | n | % |
| Included Patients | 793 |  | 132 |  |
| Included Images | 1501 |  | 239 |  |
| Patient Demographics |  |  |  |  |
| Mean age (years) | 65.00 |  | 67.83 |  |
| Age SD (years) | 14.0 |  | 13.84 |  |
| Male Proportion | 486 | 61.4 | 82 | 62.1 |
| DME Grade (Images) |  |  |  |  |
| NORMAL | 943 | 62.8 | 134 | 56.1 |
| NCI-DME | 155 | 10.3 | 32 | 13.5 |
| CI-DME | 403 | 26.8 | 73 | 30.5 |

SD, standard deviation; DME, diabetic macular oedema; NCI, non-centre-involving; CI, centre-involving.

### Detection of DME

The results for the detection of DME are presented in Table 3. This includes the results from the cross-validation training and then compares the trained algorithm results to the gradings from the first ophthalmologist and the second ophthalmologist. The results are also included when the algorithm is tested with the external dataset. The confusion matrices and the AUC curves for the grading completed by the first and second ophthalmologist and for the external dataset are included as supplemental Figures 1 and 2.

**Table 3:**
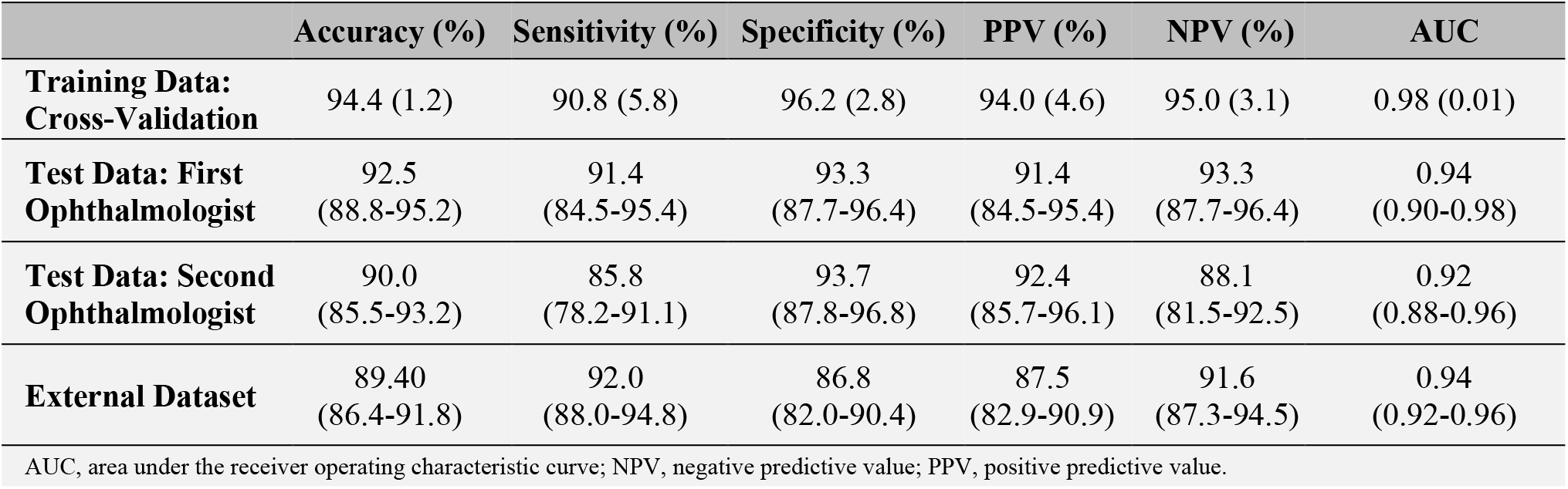
Results for DME detection using the dataset gradings. Values reported as mean (SD) for the cross-validation data The trained algorithm is then compared to the ophthalmologist gradings for the test dataset and to the external dataset, presented with 95% confidence intervals.

|  | Accuracy (%) | Sensitivity (%) | Specificity (%) | PPV (%) | NPV (%) | AUC |
| --- | --- | --- | --- | --- | --- | --- |
| **Training Data: Cross-Validation** | 94.4 (1.2) | 90.8 (5.8) | 96.2 (2.8) | 94.0 (4.6) | 95.0 (3.1) | 0.98 (0.01) |
| **Test Data: First Ophthalmologist** | 92.5 (88.8-95.2) | 91.4 (84.5-95.4) | 93.3 (87.7-96.4) | 91.4 (84.5-95.4) | 93.3 (87.7-96.4) | 0.94 (0.90-0.98) |
| **Test Data: Second Ophthalmologist** | 90.0 (85.5-93.2) | 85.8 (78.2-91.1) | 93.7 (87.8-96.8) | 92.4 (85.7-96.1) | 88.1 (81.5-92.5) | 0.92 (0.88-0.96) |
| **External Dataset** | 89.40 (86.4-91.8) | 92.0 (88.0-94.8) | 86.8 (82.0-90.4) | 87.5 (82.9-90.9) | 91.6 (87.3-94.5) | 0.94 (0.92-0.96) |

AUC, area under the receiver operating characteristic curve; NPV, negative predictive value; PPV, positive predictive value.

### Classification of DME

The mean accuracy overall for the classification of the DME type when evaluated for the 5-fold cross-validation was 91.60% (SD = 0.84%). When the algorithm for DME classification was evaluated using the test dataset graded by the first ophthalmologist, the overall accuracy of the algorithm was 89.12%. When compared to the second ophthalmologist, the accuracy of the algorithm was 84.94%. The AUC values per class are presented in Table 4 with other metrics for the cross-validation and test data. The confusion matrices and the AUC curves comparing to the DME classification algorithm are included as supplemental Figures 3 and 4.

**Table 4:**
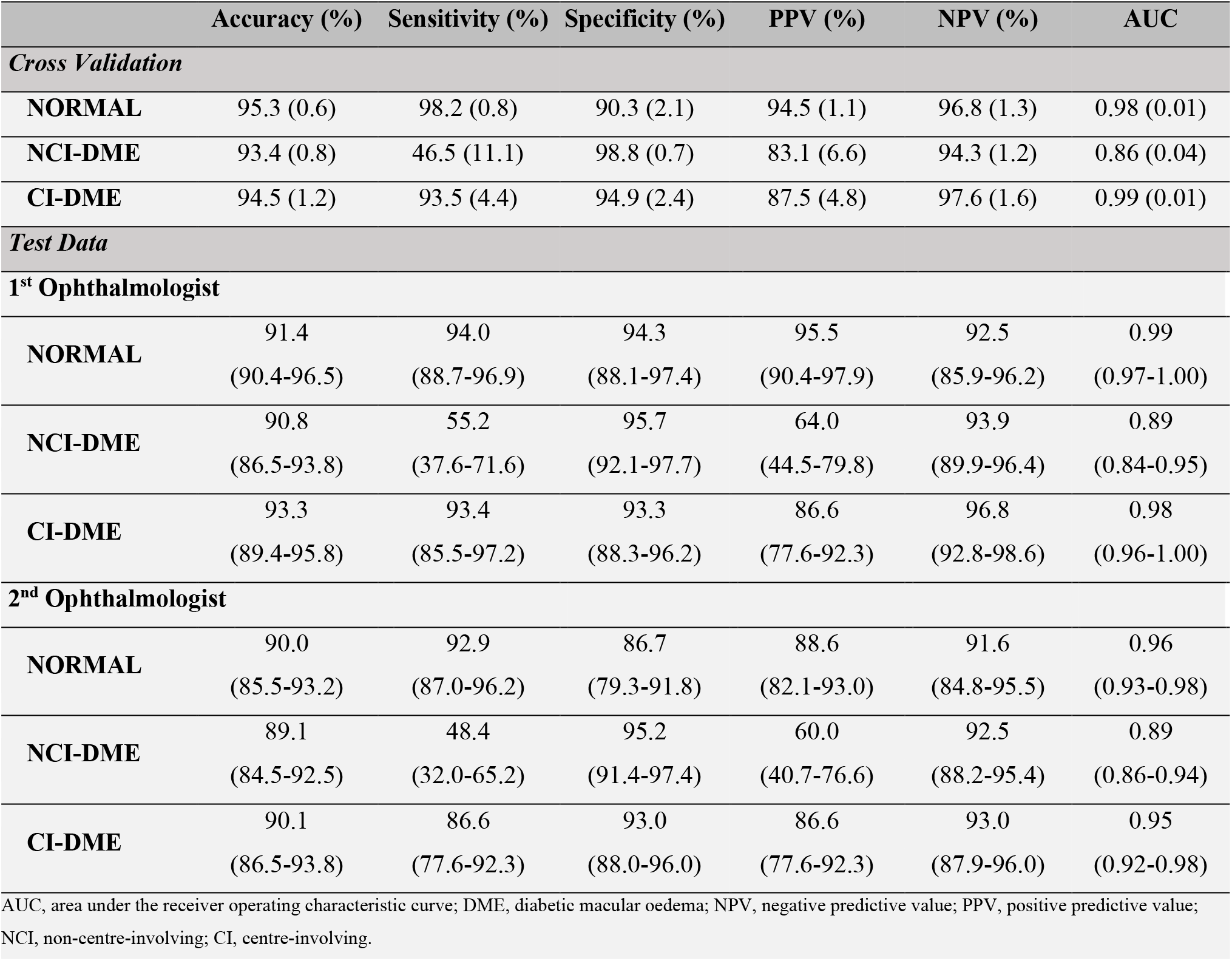
Results for the DME classification using the dataset grading. Values reported as mean (SD) for the cross-validation data. The trained algorithm is then compared using the test dataset as graded by both the first and second ophthalmologist with values presented with 95% confidence intervals.

|  | Accuracy (%) | Sensitivity (%) | Specificity (%) | PPV (%) | NPV (%) | AUC |
| --- | --- | --- | --- | --- | --- | --- |
| **Cross Validation** |  |  |  |  |  |  |
| **NORMAL** | 95.3 (0.6) | 98.2 (0.8) | 90.3 (2.1) | 94.5 (1.1) | 96.8 (1.3) | 0.98 (0.01) |
| **NCI-DME** | 93.4 (0.8) | 46.5 (11.1) | 98.8 (0.7) | 83.1 (6.6) | 94.3 (1.2) | 0.86 (0.04) |
| **CI-DME** | 94.5 (1.2) | 93.5 (4.4) | 94.9 (2.4) | 87.5 (4.8) | 97.6 (1.6) | 0.99 (0.01) |
| **Test Data** |  |  |  |  |  |  |
| **1<sup>st</sup> Ophthalmologist** |  |  |  |  |  |  |
| **NORMAL** | 91.4 (90.4-96.5) | 94.0 (88.7-96.9) | 94.3 (88.1-97.4) | 95.5 (90.4-97.9) | 92.5 (85.9-96.2) | 0.99 (0.97-1.00) |
| **NCI-DME** | 90.8 (86.5-93.8) | 55.2 (37.6-71.6) | 95.7 (92.1-97.7) | 64.0 (44.5-79.8) | 93.9 (89.9-96.4) | 0.89 (0.84-0.95) |
| **CI-DME** | 93.3 (89.4-95.8) | 93.4 (85.5-97.2) | 93.3 (88.3-96.2) | 86.6 (77.6-92.3) | 96.8 (92.8-98.6) | 0.98 (0.96-1.00) |
| **2<sup>nd</sup> Ophthalmologist** |  |  |  |  |  |  |
| **NORMAL** | 90.0 (85.5-93.2) | 92.9 (87.0-96.2) | 86.7 (79.3-91.8) | 88.6 (82.1-93.0) | 91.6 (84.8-95.5) | 0.96 (0.93-0.98) |
| **NCI-DME** | 89.1 (84.5-92.5) | 48.4 (32.0-65.2) | 95.2 (91.4-97.4) | 60.0 (40.7-76.6) | 92.5 (88.2-95.4) | 0.89 (0.86-0.94) |
| **CI-DME** | 90.1 (86.5-93.8) | 86.6 (77.6-92.3) | 93.0 (88.0-96.0) | 86.6 (77.6-92.3) | 93.0 (87.9-96.0) | 0.95 (0.92-0.98) |

AUC, area under the receiver operating characteristic curve; DME, diabetic macular oedema; NPV, negative predictive value; PPV, positive predictive value; NCI, non-centre-involving; CI, centre-involving.

## DISCUSSION

The algorithm in this study demonstrates excellent performance for the detection of DME on the cross-validated training and similar performance on the test data when compared to the gradings by the first ophthalmologist, achieving an accuracy of 92.5% and an AUC of 0.94. When compared to the second ophthalmologist, the accuracy of the algorithm is slightly reduced at 90.0% with an AUC of 0.92 although the specificity and PPV is similar in both cases. Given that the first ophthalmologist produced the gradings for the training dataset, this is likely a source of the slight difference in accuracy. The similar specificity however suggests the second ophthalmologist may have a slightly lower threshold for diagnosing DME.

The DME detection algorithm also showed favourable performance when compared to the external dataset, with an AUC of 0.94, matching the test dataset performance for the first ophthalmologist. The previous study using this dataset did achieve a higher AUC of 0.99 for binary classification using this as a test dataset, but not as an external dataset and multiple images from the same patient were used in this study.[11] The images used in this dataset also covered a wider section of the retina and could have been interpreted differently when re-sized for input in this study. There can also be slight ethnic difference between populations and has been suggested this could cause slight differences in algorithm performance when using OCT scans.[27] However, while these slight differences might contribute to variance between algorithms when tested on different populations, this study overall has demonstrated reliable performance for the detection of DME in an Irish population, where the use of DL for OCT image analysis has not previously been tested.

The DME classification algorithm in this study has also shown promising performance. The cross-validation training shows high performance, particularly for the detection of the normal and CI-DME class. When compared to the first ophthalmologist using the test data, the algorithm performs favourably for the detection of the normal class (AUC = 0.99) and the CI-DME class (AUC = 0.98), and these results are similar when compared to the second ophthalmologist using the test data for normal (AUC = 0.96) and for CI-DME (AUC = 0.95). The NCI-DME class is less well detected when compared to both the first ophthalmologist (AUC = 0.89) and the second ophthalmologist (AUC = 0.89). However, it can be noted that the NCI-DME class also has the lowest number of samples, which could affect the performance, particularly for early or mild DME. It can also be noted that as per the DME detection phase, variability between the ophthalmologist gradings may affect the algorithm predictions, and it can be noted that the algorithm performs slightly more favourably when compared to the first ophthalmologist gradings. This situation is similar for the testing of DME detection and could be improved in future work by having an increased number of graders involved in the training stage and using an increased number of images, particularly for the NCI-DME class. Furthermore, discussing difference in assigned grades would establish a more robust gold standard, and reduce the potential for any bias in grading.

Overall, the algorithms in this study have demonstrated the ability to classify DME, and there are notably low rates of error for the CI-DME class, which would be ideal for use in screening applications. In the previous similar study by Tang et al., they achieved an AUC of 0.958 with their primary dataset, and AUC results between 0.936 and 0.956 for their external datasets using images from the Heidelberg Spectralis machine for the detection of DME.[27] They achieved an AUC of 0.951 for the distinguishing of CI-DME versus NCI-DME with their primary dataset, and AUC scores between 0.899 and 0.934 for their external datasets. Thus, the overall AUC scores achieved in this study are similar and even though the accuracy scores are higher for the study completed by Tang et al., the datasets used have many images taken from the same patients, unlike this study which had no patient duplication and may have more heterogenous data. Despite the images also being classified in a slightly different manner in the previous study, this work supports the previous finding that an automated classification of DME level from OCT scans is feasible.

This has practical implications and can be used to direct treatment decisions and prevent blindness due to DME, which is relevant as the number of patients with DM continues to grow worldwide.[1] The ICO has guidelines for the management of DME based on the location on the retina,[2] and the Scottish diabetic retinal screening service already use DME levels in retinal photographs to direct treatment or referral.[41] While macular thickening may not always be associated with a change in the patient’s vision,[28] reduced visual acuity is associated with a higher prevalence of DME,[4] and thus combining visual acuity with the level DME as has been previously suggested may represent a suitable implementation of automated DME severity classification in screening programmes.[2,31] A reduction in vision in conjunction with the presence of severe DME as per the proposed grading scale should indicate the most urgent referral, and using such software in a screening service could expedite treatment for those who need the earliest treatment.

There are several limitations to consider for this study. There was a limited variety in image grading, and the gold standard for the training and validation images was set by a single ophthalmologist. It has previously been noted that inter-grader discrepancy can arise in the assessment of DME [42], and while we have shown that considerable agreement does exist between the graders for the test set, future work should utilise multiple graders for the training and validation data also, with discussion on any disagreement. Taking such steps with the grading would reduce the potential for grading bias, making the algorithm more generalisable in the future. There is a population bias as the images were only taken from a hospital ophthalmology clinic, and the amount of data was also limited in this study due to time constraints. This is most notable for the NCI-DME class, where the number of samples was limited and performance was reduced. Increasing the amount of data for this class in particular could increase future performance. While an automated approach could be used for the detection of the fovea,[43] there could be signs of DME elsewhere in the eye and only images at the foveal level were included. Furthermore, only a single model of OCT machine was used, which is limiting as characteristics can vary between devices and future work will incorporate testing with other OCT machines to ensure wider clinical applicability, possibly with adjustments per device if required. The exclusion criteria were applied to simplify the approach, but this does not reflect the full extent of retinal disease that would be encountered in a real-world screening service as other retinal pathologies can often exist with DME, and this would need to be included in future work to ensure clinical applicability. It is also worth noting that the algorithms in this test overall show lower sensitivity compared to specificity for the test data for DME detection and classification and could lead to increased missed cases over un-needed referrals. This may be less ideal depending on the requirements of a given screening programme, but further work could address this potentially using more sophisticated DL techniques. Alternatively, an adjustment to the algorithm probability threshold may reduce false negatives at the expense of increasing false positives but may be acceptable in a screening setting to avoid missed cases, which may be most applicable to the NCI-DME class where the performance was reduced. Further testing could also be carried out against other deep learning implementations for OCT analysis, and this should be considered in further work. As clinicians and patients alike expect that AI systems should be reliable,[44] it is also important to consider how algorithms can provide a rationale for their prediction. This can be achieved by highlighting the most influential area on a scan,[44] and could also be implemented in future work.

## CONCLUSIONS

This study has shown that DME severity can be classified from OCT scan images using DL using a recognised scale for DME. This supports the findings of previous similar work and adds to the evidence for the automated classification of DME severity, including in an Irish population. Such a system and the utilised grading scale has clinical significance as it could speed up referral times for those in need of urgent treatment without compromising patient safety, and to effectively use limited resources. Further work could likely improve the classification performance.

## Data Availability

Data used in this study are available on reasonable request for further development in this area. Project code is also available on request and is also available online with the results data.

https://github.com/cbreathnach/DME-Detection

## ADDITIONAL STATEMENTS

### Ethics

Ethics approval for this study was obtained from the Clinical Research Ethics Committee at Galway University Hospitals with reference number C.A. 2831. Informed consent was not required from patient due to the retrospective nature of the study, and the use of fully anonymised images. This study was not registered with any other body.

### Contributors

DOK, FH and CB proposed the study. DOK, FH, CB and AS contributed to the study design and methods, including the analysis. CB and RH collected and organised the data. FH and DT graded the images. CB wrote the software. CB prepared the original draft manuscript. DOK, FH, DT, RH, AS reviewed the manuscript. All authors have read and agreed to the final version of the manuscript.

### Data availability statement

Data are available on reasonable request. Project code with the prediction data is referenced in the supplemental material and are available online.

