## supplemental material for "Classifying the severity of diabetic macular oedema from optical coherence tomography scans using deep learning: a feasibility study"

| N | Layer | Output Shape |
|---|---|---|
| 1 | Input & Rescaling | 256 x 256 x 1 |
| 2 | Conv2D (kernel size = 3 x 3) | 256 x 256 x 16 |
| 3 | MaxPooling2D (pool size = 2 x 2) | 128 x 128 x 16 |
| 4 | Conv2D (kernel size = 3 x 3) | 128 x 128 x 32 |
| 5 | MaxPooling2D (pool size = 2 x 2) | 64 x 64 x 32 |
| 6 | Conv2D (kernel size = 3 x 3) | 64 x 64 x 64 |
| 7 | MaxPooling2D (pool size = 2 x 2) | 32 x 32 x 64 |
| 8 | Conv2D (kernel size = 3 x 3) | 32 x 32 x 128 |
| 9 | MaxPooling2D (pool size = 2 x 2) | 16 x 16 x 128 |
| 10 | Conv2D (kernel size = 3 x 3) | 16 x 16 x 256 |
| 11 | BatchNormalization | 16 x 16 x 256 |
| 12 | MaxPooling2D (pool size = 2 x 2) | 8 x 8 x 256 |
| 13 | Dense | 8 x 8 x 256 |
| 14 | Dropout (rate = 0.2) | 8 x 8 x 256 |
| 15 | Flattening | 16384 |
| 16 | Dense | 128 |
| 17 | Dense | Number of Classes |

Supplemental Table 1: CNN structure. Model output as probability per class.

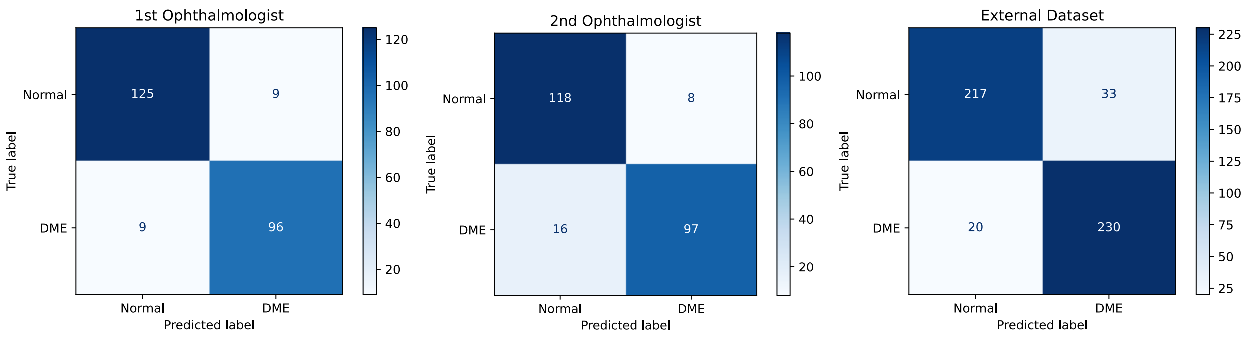


Supplemental Figure 1: Confusion matrices for the test data for DME detection compared to the first ophthalmologist (left), compared to the second ophthalmologist (middle) and compared to the external dataset gradings (right).

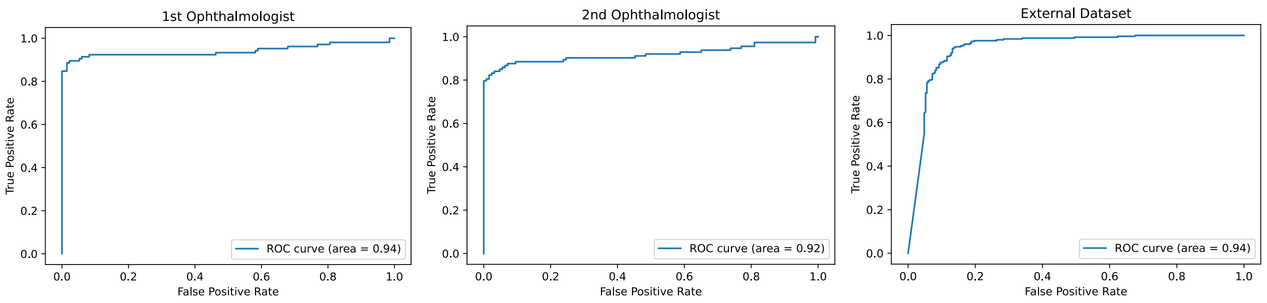


Supplemental Figure 2: AUC curves for the test data for DME detection compared to the first ophthalmologist (left), compared to the second ophthalmologist (middle) and compared to the external dataset (right).

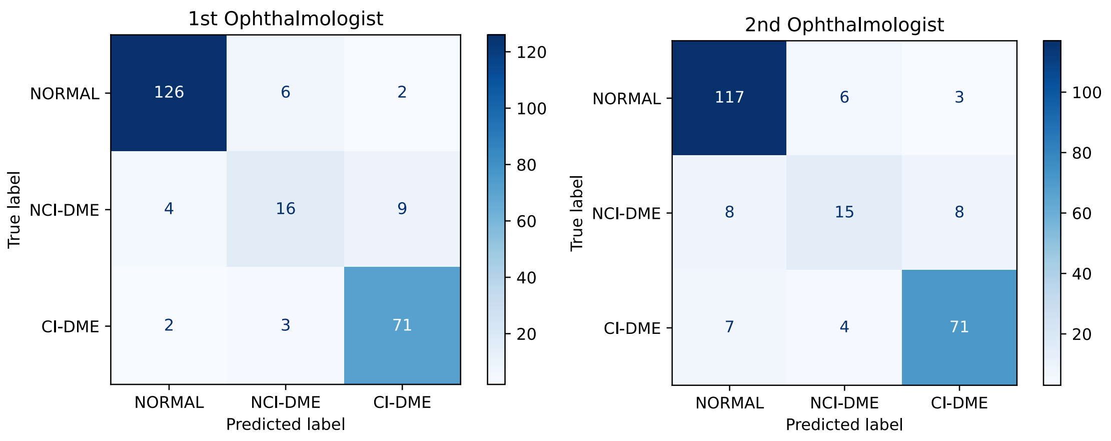


Supplemental Figure 3: AUC curves for the test data for DME classification compared to the first ophthalmologist (left) versus second ophthalmologist (right).

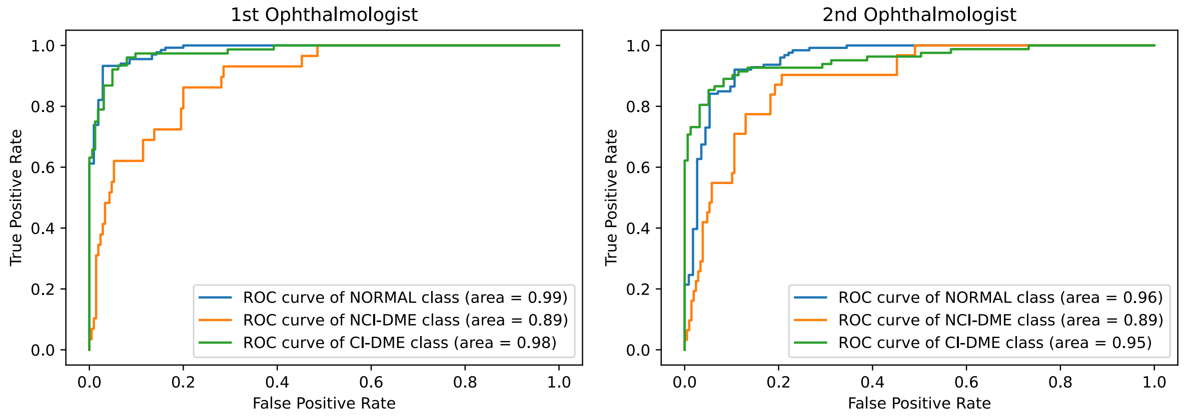


Supplemental Figure 4: AUC curves for the test data for DME classification compared to the first ophthalmologist (left) versus second ophthalmologist (right).

**Study Protocol:** The study was carried out as outlined in the primary manuscript, and an additional study protocol was not prepared.

**Code:** The code for this project is available online on Github: https://github.com/cbreathnach/DME-Detection
